# Streamlining Intersectoral Provision of Real-World Health Data: A Service Platform for Improved Clinical Research and Patient Care

**DOI:** 10.1101/2024.01.29.24301922

**Authors:** Katja Hoffmann, Igor Nesterow, Yuan Peng, Elisa Henke, Daniela Barnett, Cigdem Klengel, Mirko Gruhl, Martin Bartos, Frank Nüßler, Richard Gebler, Sophia Grummt, Anne Seim, Franziska Bathelt, Ines Reinecke, Markus Wolfien, Jens Weidner, Martin Sedlmayr

## Abstract

**Introduction:** Obtaining real-world data from routine clinical care is of growing interest for scientific research and personalized medicine. Despite the abundance of medical data across various facilities — including hospitals, outpatient clinics, and physician practices — the intersectoral exchange of information remains largely hindered due to differences in data structure, content, and adherence to data protection regulations. In response to this challenge, the Medical Informatics Initiative (MII) was launched in Germany, focusing initially on university hospitals to foster the exchange and utilization of real-world data through the development of standardized methods and tools, including the creation of a common core dataset. Our aim, as part of the Medical Informatics Research Hub in Saxony (MiHUBx), is to extend the MII concepts to non-university healthcare providers in a more seamless manner to enable the exchange of real-world data among intersectoral medical sites.

**Methods:** We investigated what services are needed to facilitate the provision of harmonized real-world data for cross-site research. On this basis, we designed a Service Platform Prototype that hosts services for data harmonization, adhering to the globally recognized Health Level 7 (HL7) Fast Healthcare Interoperability Resources (FHIR) international standard communication format and the Observational Medical Outcomes Partnership (OMOP) Common Data Model (CDM). Leveraging these standards, we implemented additional services facilitating data utilization, exchange and analysis. Throughout the development phase, we collaborated with an interdisciplinary team of experts from the fields of system administration, software engineering and technology acceptance to ensure that the solution is sustainable and reusable in the long term.

**Results:** We have developed the pre-built packages “ResearchData-to-FHIR”, “FHIR-to-OMOP” and “Addons”, which provide the services for data harmonization and provision of project-related real-world data in both the FHIR MII Core dataset format (CDS) and the OMOP CDM format as well as utilization and a Service Platform Prototype to streamline data management and use.

**Conclusion:** Our development shows a possible approach to extend the MII concepts to non-university healthcare providers to enable cross-site research on real-world data. Our Service Platform Prototype can thus pave the way for intersectoral data sharing, federated analysis, and provision of SMART-on-FHIR applications to support clinical decision making.

## 1. Introduction

For scientific research, there is a high interest in using data from routine clinical care, so-called real-world data (RWD) (1–4). Although large amounts of RWD are available in various institutions, such as hospitals, outpatient clinics, and physician practices, the intersectoral data exchange between sites is hindered by their heterogeneity in terms of structure, content, and compliance with data protection regulations (2,5). To address this challenge, the German Medical Informatics Initiative (MII) was launched in 2018, initially focusing on university hospitals to foster the exchange and utilization of RWD (6,7). At that time, Data Integration Centers (DIC) were established at the medical sites of the university hospitals, and standardized solutions were developed for effective data use and exchange in both healthcare and research, with a focus on interoperability and data harmonization. For example, all university hospitals in the MII defined a dataset description, the MII Core dataset (CDS) (8) using the Health Level 7 (HL7) Fast Healthcare Interoperability Resources (FHIR) international standard communication format (9). The FHIR MII CDS consists of basic modules (e.g., Person, Case, Diagnosis, Procedure, Laboratory test results, medication), and extension modules (e.g., Oncology, Pathology results, Molecular Genetics, Intensive care) (8). This forms the foundation for cross-site data exchange and the integration of third-party applications via SMART-on-FHIR technology (10).

To facilitate the data exchange of RWD for scientific purposes, the HL7 International (11) and the Observational Health Data Sciences and Informatics (OHDSI) (12,13) community have announced collaboration in 2021 (14). The OHDSI community develops the Observational Medical Outcomes Partnership (OMOP) common data model (CDM) (15), as well as tools for data quality assessment and analysis (16). Common to all standardized data formats is the need to develop individual processes for the extraction, transformation and loading (ETL) of data from different data sources, which remains a major challenge (5). Nevertheless, hospitals have succeeded in overcoming these initial hurdles and making their own RWD available in a harmonized form for research (17,18) and possibly also for patient care.

Since certain diseases, such as cardiovascular diseases, diabetes, allergies, and mental illnesses, are often not treated at university hospitals and there is insufficient data available, especially for rare diseases, it is essential to obtain RWD as well from non-university healthcare providers to ensure a comprehensive and diverse dataset for research studies. In order to make RWD available for research by non-university healthcare providers, the Medical Informatics Hub in Saxony (MiHUBx) was founded in 2021 (19). Among the project goals is to investigate whether the concepts of the MII can be transferred to non-university healthcare providers. As a first result within this project, Bathelt et al. (2022) demonstrated the possibility to utilize an existing portable and standardized infrastructure from a university hospital setting and transferred it to non-university sites to support feasibility requests for participation in multicentre studies (1). However, the work of Bathelt et al. was limited in terms of data availability, terminologies used, and harmonized data formats, so that RWD from non-university sites are still insufficiently available for research. Yet, due to limited human and economic resources and expertise in HL7 FHIR and OMOP CDM, it is hardly possible for non-university healthcare providers to develop the required services for data harmonization and provision in standardized data formats themselves. It is therefore necessary to provide the services in such a way that they can be conveniently deployed and easily used by data providers for different studies. In this paper, all tools for data harmonization, provision and management, such as databases, programs for ETL processes, analysis tools and other applications, are referred to as services.

The aim of this work is the development of pre-built packages that contain services and support the harmonization and provision of RWD in FHIR MII CDS and OMOP CDM format and thereby provide reuse potential for various projects. Another goal of this work is the development of a versatile and modular Service Platform Prototype that facilitates project administration, service management, data management and analysis. In this context, we focus on the following two research questions:

1. What services are needed to facilitate the provision of harmonized RWD for cross-site research?
2. How can the necessary services (from 1) be technically compiled so that they can be used by hospitals with few resources and limited expertise in HL7 FHIR and OMOP CDM to make harmonized RWD accessible for research studies?

## 2. Materials and methods

### 2.1. Materials for data harmonization and provision

#### 2.1.1. Compilation and integrated implementation of existing software resources

For the harmonization of RWD from source systems into the basic modules of the FHIR MII CDS version 1.0 (20), and into the OMOP CDM format (15), we have selected the following materials, as these are successfully used at the University Hospital Dresden. In addition, we investigated the applicability of other established tools. For this purpose, we used the following developed concepts of the MII and the software tools published by the MII consortium Medical Informatics in Research and Care in University Medicine (MIRACUM) (21) as MIRACOLIX Tools (22):

##### Clinical Data Repository

A Clinical Data Repository (CDR) is a database in which patient-centered healthcare data from various IT systems (e.g., electronic health record (EHR), laboratory system, biobank) are stored in a site-specific data model.

##### FHIR Server BLAZE

A FHIR Server is a software solution that stores and manages FHIR resources. It acts as a bridge connecting healthcare applications and systems, allowing them to exchange patient information in a consistent and structured format or to answer population-wide aggregate queries quickly. The FHIR Server BLAZE (23) was initially developed within the German Biobank Alliance project (24), aimed at high-throughput performance (25). Blaze comes with a built-in feature to authenticate requests against an OpenID Connect provider.

##### MIRACUM FHIR Gateway

The MIRACUM FHIR Gateway (26) is a PostgreSQL database with a table for storing FHIR Resources, which are represented in JSON format. It serves as a temporary storage.

##### ETL Process DWH-TO-FHIR

The ETL Process DWH-TO-FHIR extracts research data, which is provided as database views, from the data warehouse (DWH) of the site, constructs FHIR resources according to the FHIR MII CDS structure definition v1.0 and loads them into a *MIRACUM FHIR Gateway*. The application DWH-TO-FHIR provides Basic Authentication (27) to enforce access controls to the FHIR resources of the FHIR server.

##### ETL Process ROTATOR

The ROTATOR (f**RO**m ga**T**ew**A**y **TO** se**R**ver) is an application that reads FHIR resources from the MIRACUM FHIR Gateway and loads them onto a FHIR Server, such as BLAZE.

##### OHDSI Tools

The OHDSI Tools are provided as Docker containers by Gruhl et al. (28). The basis of the OHDSI Tools is the OMOP CDM PostgreSQL database, which is divided into the standardized OMOP CDM data tables v5.3 and the OMOP CDM standardized vocabularies (state February 2023), such as SNOMED CT (Systematized Nomenclature of Medicine -Clinical Terms), ICD-10-GM (International Classification of Diseases, German Modification) and OPS (“Operationenund Prozedurenschlüssel”, surgery and procedure key). Furthermore, the OHDSI Tools by Gruhl et al. (28) includes additional dockerized tools for analyzing research data in OMOP CDM: 1) the R-based application ACHILLES that can be used for data characterization and visualization, 2) the web-based application ATLAS that can be used for cohort definition and scientific analysis, and 3) the R-based application Data Quality Dashboard (DQD) that can be used for data quality analysis.

##### ETL Process FHIR-TO-OMOP

The ETL Process FHIR-TO-OMOP extracts FHIR resources from a FHIR Server (e.g., the FHIR Server BLAZE) or from the MIRACUM FHIR Gateway, transforms them into the standardized format of OMOP CDM and loads them into an OMOP CDM database (29,30). The application provides HTTP Basic Authentication (27) to enforce access controls to the FHIR resources of the FHIR server.

##### TRANSITION Database

The TRANSITION Database is a relational database that was originally developed for the semantic mapping of system-specific documented diagnoses to Orpha codes for rare diseases (23). It is deployed as a PostgreSQL database v14 (22), and offers tables with required terminology bindings (e.g., FHIR value sets), which can essentially support the semantic mapping of the RWD to required code systems. For example, the semantic mapping of the gender (e.g. value “1” for female) to a FHIR ValueSet (e.g. code “female” (24)) can be achieved via the TRANSITION Database. As an example of the database structure, the table for the semantic mapping of the vital parameters can be found as a csv file in the Supplementary Materials (S1 file).

##### Keycloak

Since medical data needs to be protected in an enhanced manner, the Keycloak server offers significant advantages (e.g. centralized Identity Management, Customizable Authentication Flows) for the medical domain, particularly in terms of securing sensitive patient data, ensuring compliance with healthcare regulations, and facilitating interoperability among diverse healthcare IT systems. The Authentication Server Keycloak (34) can be used as an OpenID (35), OAuth 2.0 (36), or SAML Connect provider to validate requests to the FHIR endpoints.

#### 2.1.2. Original Contributions - Expanding with Custom Software Additions

We analyzed the already established data provision pipeline at University Hospital Dresden by consulting experts in the fields of data integration, provision, protection and security. From this, we derived a generic process for data harmonization and provision. Subsequently, we identified missing materials, which we have developed and adapted. These are technically described below and further explained in the Results Section.

##### Research Data Repository

In order to be able to provide the ETL process DWH-TO-FHIR to other sites, a database structure is required which represents medical data and follows the logic of the basic modules of the FHIR MII CDS specification v1.0 (20). For this reason, we developed the Research Data Repository (RDR), deployed as a PostgreSQL database v14 (32). This repository encompasses tables for various FHIR resource types, like *Patient, Encounter, Condition, Observation, Procedure, Medication*, and *MedicationAdministration*, and is used for the cross-project and project-related storage of research data from routine clinical care (RWD).

##### Structural Mapping Guideline MII CDS

To reduce implementation time and to facilitate the specific data mapping, which must be done at each site, the Structural Mapping Guideline MII CDS is developed (S2 File).

##### The ETL process RDR-to-FHIR

To streamline the ETL process to provide RWD in FHIR MII CDS format, the sub-processes DWH-to-FHIR and ROTATOR were merged into the ETL process RDR-to-FHIR. RDR-to-FHIR loads project-related RWD from the RDR, constructs FHIR resources according to the FHIR MII CDS specification v1.0 and loads directly into the FHIR Server without buffering via the MIRACUM FHIR Gateway. To facilitate process execution, a RESTful API client was implemented to send data to the FHIR API. To enable OAuth 2.0 (36) authentication against the FHIR server BLAZE, we have added this authentication method to the application.

##### ETL Process FHIR-TO-OMOP

To enable OAuth 2.0 authentication against the FHIR server BLAZE, we added this authentication method to this application.

### 2.2. Pre-built Packages for data harmonization and provision

We have developed pre-built packages, e.g. for the transformation of RWD to FHIR, to facilitate creating, running and connecting the multiple services (see section 2.1). In close collaboration with an interdisciplinary team of software developers, database engineers as well as infrastructure, security, and usability experts, we defined specific applications and composed the services to easy-to-install, pre-configurable installation packages based on Docker v24 (37) and Docker Compose v2 (38). We made our decisions based on previous experience and preliminary works. For testing purposes, we provided a test dataset.

To facilitate data and system administration, we have included the following additional materials in our packages, which can be used optionally.

#### pgAdmin

The open-source administration and management tool pgAdmin4 (39) provides a user-friendly web interface for the efficient administration of PostgreSQL databases used for RDR, TRANSITION database, Keycloak, and OMOP CDM.

#### Portainer

The open source container administration tool Portainer community edition (40) simplifies the management of containerized applications that are used to provide all the services described in the section 2.1. Portainer provides a web interface for interacting with Docker containers (37) to create, manage, and deploy containers without the need for extensive knowledge of Docker commands.

### 2.3. Service Platform Prototype

To support research projects and system administration, we have developed a web-based Service Platform Prototype that facilitates project-related data provision for research projects. Based on experience with the design of user interfaces for interactive systems, an initial Low-Fidelity Prototype of the service platform was created to serve as a basis for discussion for a joint, interdisciplinary Graphical User Interface (GUI) design. By incorporating feedback from usability experts, this Low-Fidelity Prototype was iteratively transformed into a High-Fidelity Prototype. To this end, regular joint meetings were held with software developers and usability experts to ensure that the interaction principles for design solutions in accordance with ISO 9241-110 (41) were taken into account and implemented in the service platform interface (task appropriateness, self-descriptiveness, conformity to expectations, learnability, controllability, robustness against errors, user retention).

Our Service Platform Prototype was developed using Quarkus v3.4.3 (42). The frontend provides a form-based interface for users to specify the services and configurations needed for their project. The backend, managing the service deployment, is responsible for the instantiation of Docker containers, network setup, configuration management, and error handling. Angular v16.2.8 (43) was used for the frontend, complemented by Angular Bootstrap v16.0.0 (44). Communication between the microservices (e.g., to manage projects and Docker containers) is orchestrated by the Java Docker API v3.3.3 (45). Communication between the services is facilitated by RabbitMQ (46), a message broker that ensures reliable and real-time messaging.

## 3. Results

### 3.1. A generic data harmonization and provision process to foster data availability

We developed a generic process for the project-related provision of research data, which is shown in Figure 1 and described below.

**Figure 1.**
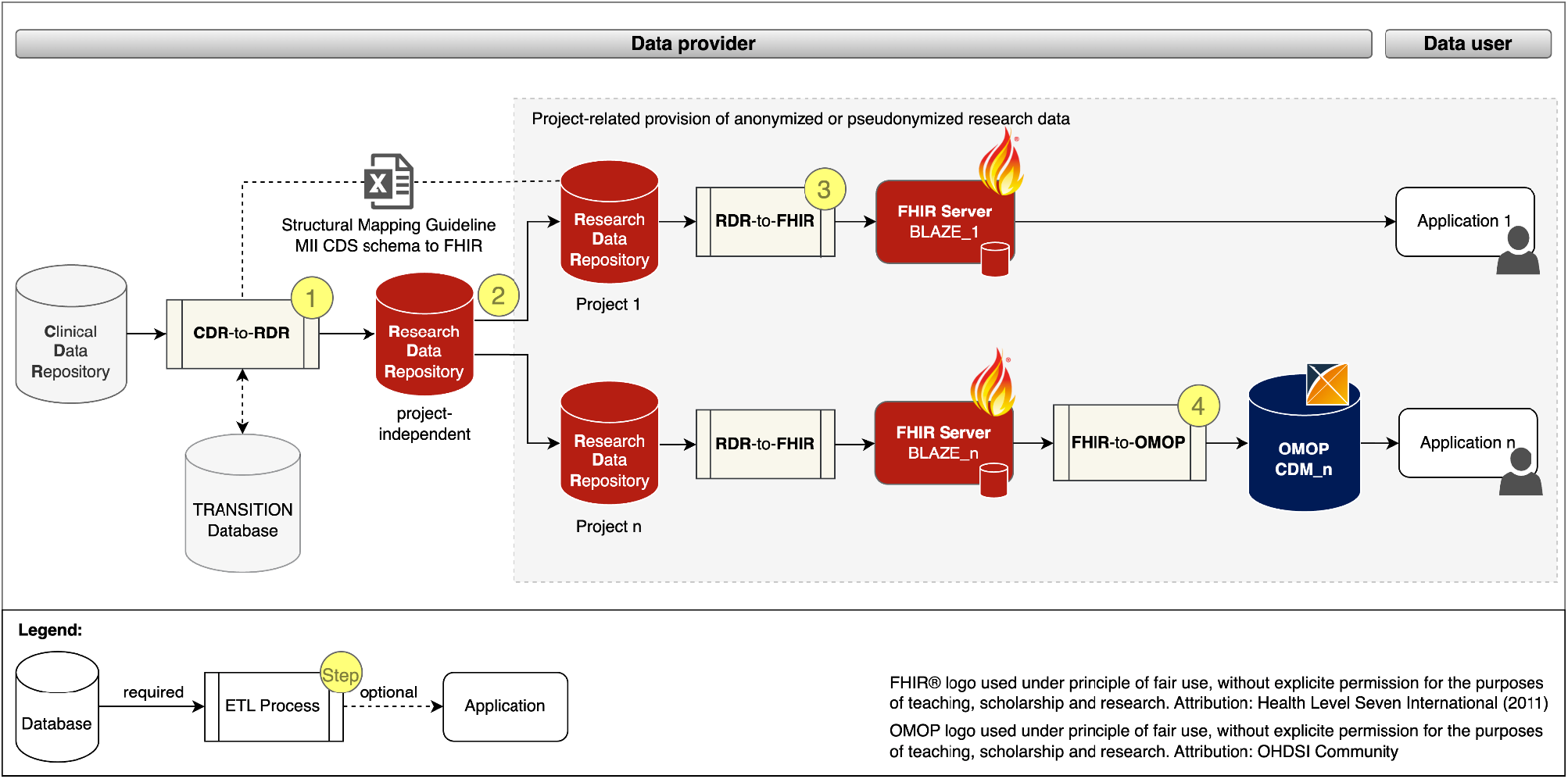
Project-related provision of research data for providers and users. Fast Healthcare Interoperability Resources (FHIR); Observational Medical Outcomes Partnership (OMOP); Common Data Model (CDM); Extract Transform Load (ETL); Clinical Data Repository (CDR); Research Data Repository (RDR): Medical Informatics Initiative (MII); Core dataset (CDS)

The data provision process begins with a project-independent and site-specific ETL pre-process CDR-TO-RDR (cf. Figure 1 (Step 1)) that extracts RWD from the CDR, transforms it with the help of the *Structural Mapping Guideline* and the *TRANSITION Database* to the data structure of the *Research Data Repository (RDR)* and loads it into a *project-independent* instance of the *RDR*.

According to the MII concepts, the organizational starting point for the provision of data for a research project is a request from the data user. After the technical and legal feasibility check, the required data for the study is requested from the *project-independent RDR* instance and masked, pseudonymized or anonymized in the subsequent process step to ensure that the research data does not allow any conclusions to be drawn about the identity of a patient (Figure 1 (Step 2)).

To provide the project-related data in FHIR MII CDS format, instances of *RDR-TO-FHIR* and the *FHIR Server BLAZE* are created and the ETL process is executed (Figure 1 (Step 3)). To provide the project-related data in the OMOP CDM format, instances of *FHIR-TO-OMOP* and the *OHDSI tools* (cf. OHDSI tools) are created and the ETL process is executed (Figure 1 (Step 4)).

### 3.2. Pre-built Packages to facilitate interoperability on the fly

We developed the following three pre-build packages: 1) ResearchData-TO-FHIR Package for converting RWD to FHIR resources (cf. red bordered box in Figure 2), 2) FHIR-TO-OMOP Package for converting the FHIR resources to the OMOP CDM format (cf. blue bordered box in Figure 2), and 3) the Addons Package for deployment of optional services to simplify database, container, and security management (cf. gray bordered box in Figure 2). Figure 2 illustrates the composition of our implemented services and the data flows.

**Figure 2.**
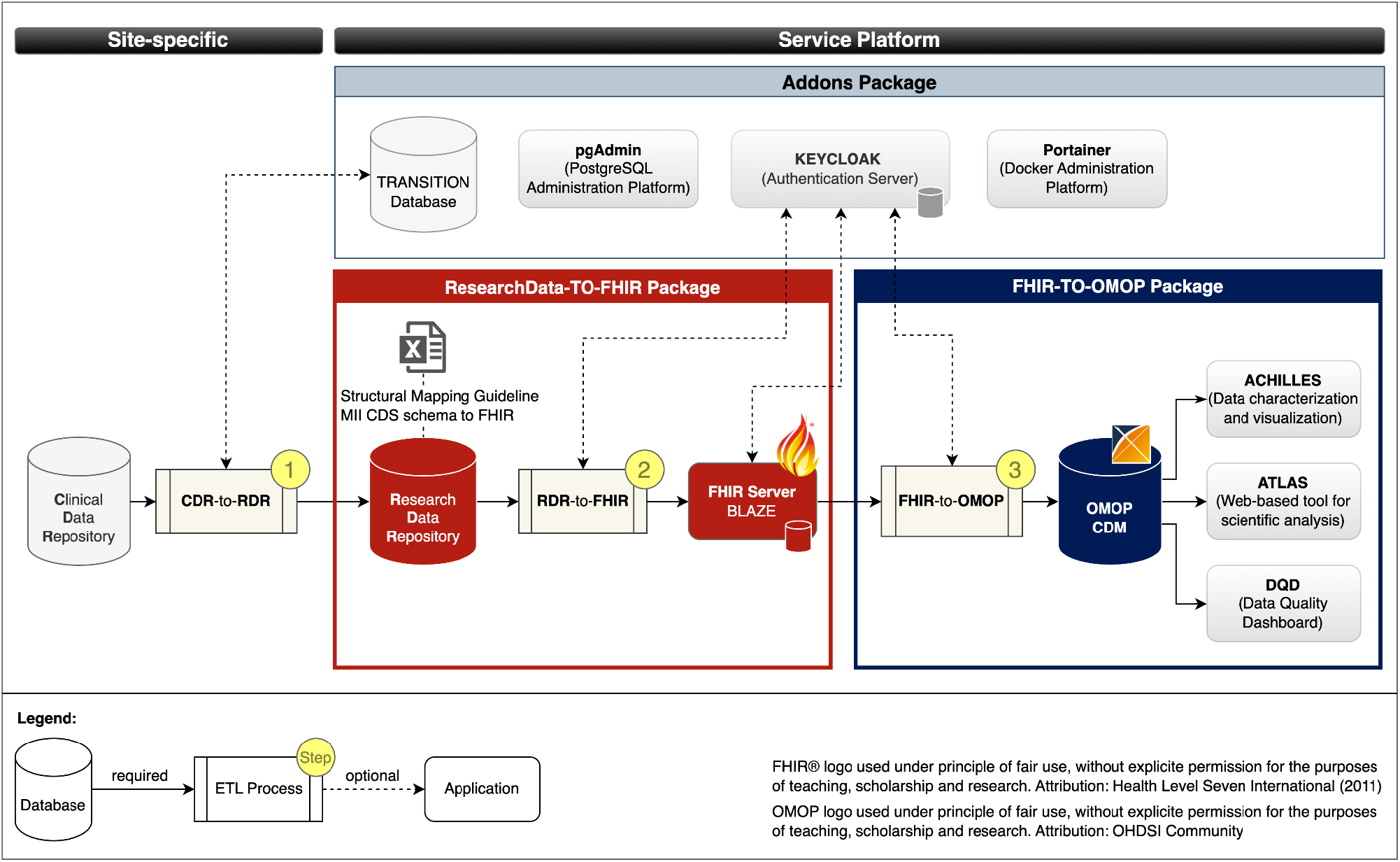
Services and data flows. Fast Healthcare Interoperability Resources (FHIR); Observational Medical Outcomes Partnership (OMOP); Common Data Model (CDM); Extract Transform Load (ETL); Clinical Data Repository (CDR); Research Data Repository (RDR): Medical Informatics Initiative (MII); Core dataset (CDS)

#### ResearchData-TO-FHIR Package

The ResearchData-TO-FHIR Package v2.2.0 (36) provides a Docker compose specification that allows to automatically retrieve the images of the *RDR*, the *FHIR server BLAZE*, and the *RDR-to-FHIR* (cf. section 2.1 and red bordered box in Figure 3), creates Docker containers as instances of the images that can be used to provide project-related research data in FHIR MII CDS format. For testing purposes, the package provides a test dataset. The package also includes the Structural Mapping Guideline (cf. Research Data Repository). By adjusting environment variables, the installation is customizable.

**Figure 3.**
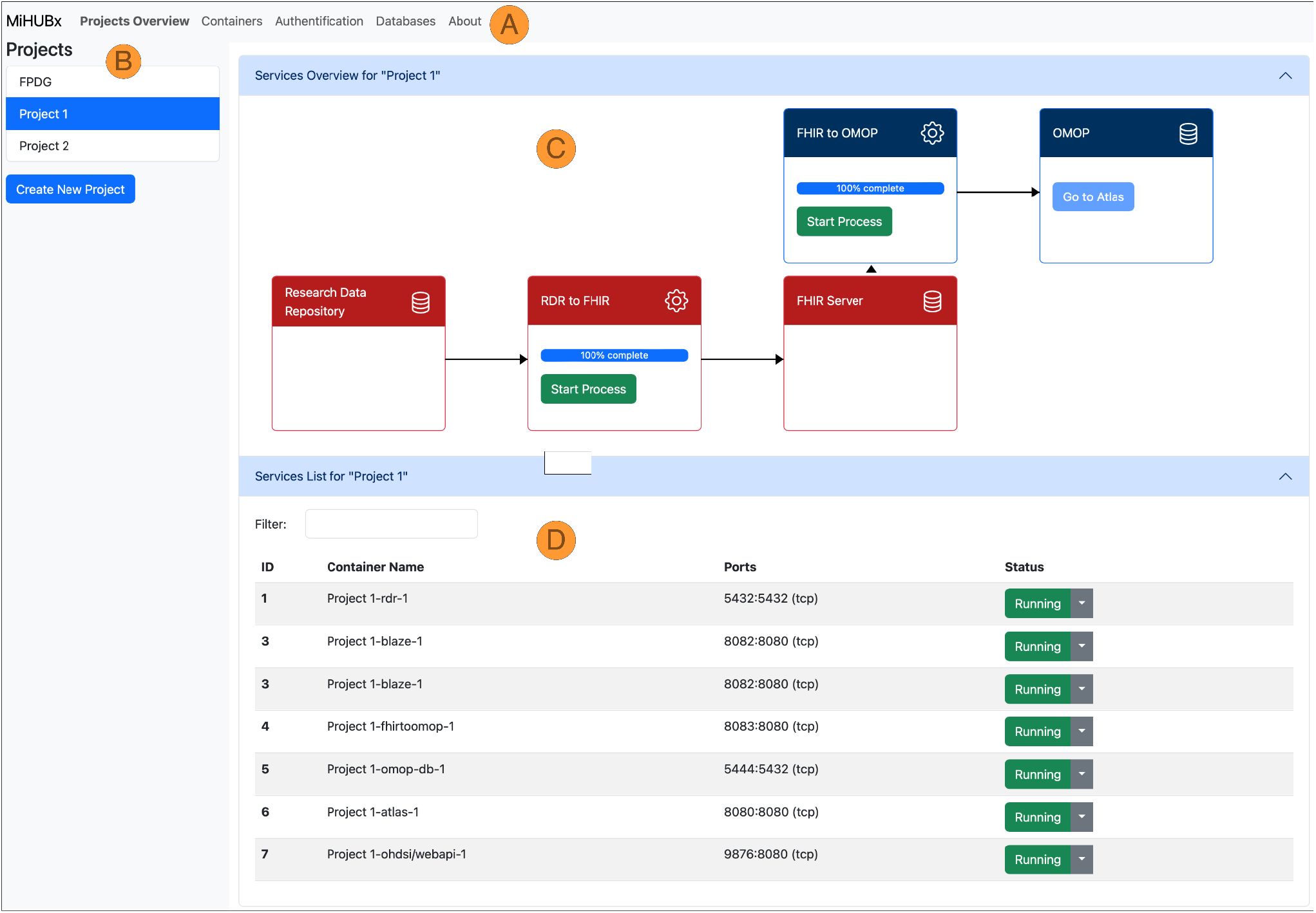
Schematic representation of the Service Platform Prototype. Fast Healthcare Interoperability Resources (FHIR); Observational Medical Outcomes Partnership (OMOP); Research Data Repository (RDR); Identifier (ID); Medical Informatics Hub in Saxony (MiHUBx)

#### FHIR-TO-OMOP Package

The FHIR-TO-OMOP Package v1.1.0 (37) provides a Docker compose specification that allows to automatically retrieve the images of the *OHDSI Tools* and the *FHIR-TO-OMOP* (cf. section 2.1 and blue bordered box in Figure 3), creates Docker containers as instances of the images that can be used to provide project-related research data in OMOP CDM format. Through environment variables, the installation is customizable and the synthetic dataset from the ResearchData-TO-FHIR package can be used for testing purposes.

#### Addons Package

The Addons Package v1.0.0 (38) provides Docker compose specifications that allow to optionally deploy the PostgreSQL administration platform *pgAdmin*, the *TRANSITION database*, the container administration tool *Portainer* and the Authentication server *Keycloak* with a demo configuration for protecting the FHIR Server Blaze (cf. ResearchData-TO-FHIR Package).

The source code of the ResearchData-TO-FHIR package v2.2.1 (47), the FHIR-TO-OMOP package v1.1.0 (48) and the Addons package v1.0.0 (49) includes instructions for installation and usage as well as for further developments.

### 3.3. Service Platform Prototype to enable an easy to use and modular infrastructure

While the pre-built packages and their services, which are integrated into the Service Platform Prototype, were described in 3.2, the Service Platform Prototype v1.0.0 (50) itself is described below.

We implemented two key services: 1) the Project Management Service and 2) the Container Management Service. The Project Management Service allows researchers to create and manage their research projects. The Container Management Service automates the installation, operation and administration of services for each research project, such as the launch of ETL-processes and the display of technical details, such as unique identifiers, Docker names, assigned communication ports or operational status, to inform users about the current activity and availability of services.

The frontend layout contains a navigation bar (Figure 3, A), a sidebar (Figure 3, B) and a main content area (Figure 3, C+D). The navigation bar offers links to subpages that provide services and further information on database administration (i.e. via pgAdmin), authentication (i.e. Keycloak) and container administration (i.e. Portainer). The research projects are listed in the sidebar and new projects can be created via clicking on the respective button. The main content area is divided horizontally. The upper area displays the project-related services and offers functions for use and management, such as starting and stopping the Docker containers, starting the ETL processes, accessing the web frontend of the services and receiving further information (Figure 3, C). The project-related containers are listed in the lower area, where further technical details are displayed, e.g. information on Docker containers, images, communication ports and operating status (Figure 3, D).

The demo server, as an instance of the Service Platform Prototype, is available at https://tudresden.de/med/demoserver. This repository (50) also contains the developer documentation, including initial installation instructions.

## 4. Discussion

Our aim was to determine which services and processes are required to enable the provision of RWD for cross-site research and patient care, and how the services can be made available and usable to non-university healthcare providers with limited resources in a low-threshold manner. In particular, we determined which services are required for the harmonization of healthcare data into HL7 FHIR standard-based MII CDS format for intersectoral exchange, and to the OMOP CDM format for national or even international cross-site research. From this, we derived a process for the project-related provision of RWD based on the already established data provision pipelines of the university sites (21) in Germany. On this basis, we developed the pre-built packages *ResearchData-TO-FHIR, FHIR-TO-OMOP*, and *Addons* for data harmonization and provision to facilitate interoperability on the fly. To streamline the use of the particular services, especially for non-university healthcare providers, we developed a versatile and modular Service Platform Prototype that demonstrates the administration of research projects based on RWD.

To the best of our knowledge, our work is the first that shows how services for data harmonization, provision, and analytics can be provided to non-university healthcare providers in a low-threshold manner. The proposed Pre-built Packages and Service Platform Prototype streamline the process of setting up research project environments and reduce the time and technical expertise required to provide RWD for research studies and feasibility inquiries, such as those conducted by the German Portal for Medical Research (Deutsches Forschungsdatenportal für Gesundheit, FDPG) (18,51). Even though admission to the FDPG is currently only possible for sites that participate in the MII (mainly universities), our work is also highly relevant for non-university service providers, as feasibility studies can also be carried out using the OHDSI tools. We also consider it likely that our work may be of use to the European Health Data Space (EHDS) (52) in the future, as our platform can be extended with appropriate services for data harmonization.

Although our work is a big step towards intersectoral provision of RWD, it has some limitations due to the following factors:

1. Healthcare providers use site-specific clinical information systems to store RWD, which have limited standardized interfaces for the data exchange. Therefore, the RWD from possibly multiple data sources must be harmonized into the data format of our provided Research Data Repository in a pre-processing step by the sites themselves, which can be a challenge depending on the type and scope of data storage, the available human and economic resources and the knowledge of medical informatics. The legally binding interface specifications currently developed and established in Germany, such as the HL7 FHIR standard-based *Information technology systems in hospitals* (Informationstechnische Systeme im Krankenhäusern, ISIK) (53) and *Medical Information Objects* (Medizinische Informationsobjekte, MIO) (54), could provide a remedy here, as services based on these standards could be developed and made available via our platform to automatically convert the RWD into the FHIR MII CDS format for research purposes.
2. The FHIR MII CDS v1.0, established as the standard for data exchange within the MII, presents certain limitations, particularly in its dataset specifications for specific medical fields like oncology and ophthalmology. Although plans are in place to refine profiles for oncology in the upcoming FHIR MII CDS v2, there remains a need to develop more appropriate dataset specifications for specific medical domains. These improvements are crucial for effectively incorporating such specifications into similar service platforms, especially in key areas like *Observations, Imaging Studies, Diagnostic Reports, Procedures*, and *Medication Administrations*.
3. In addition, the services/applications cannot yet be installed “out of the box” via the frontend. In order to further minimize the technical hurdle, this is an important goal for the future.

Despite the limitations, our Pre-built Packages together with the Service Platform Prototype can already be used to provide data for specific research projects in a time-saving manner. We believe that our research represents a significant contribution in research data management, offering an efficient, user-friendly, and reproducible way to establish project-specific data provision pipelines. This may make our work interesting not only for non-university service providers, but also for university sites. In addition, our Service Platform Prototype can serve as a foundation for third-party applications, e.g. based on SMART-on-FHIR, which can be used not only for research but also for patient care.

Next, we will implement the Service System Platform at two non-university hospitals in Germany as part of a pilot study. In order to achieve a high level of acceptance among end users, we will test its functionality and usability. As part of the roll-out, an accompanying acceptance analysis in the form of an observation protocol and in-depth face-to-face interviews will be conducted to ensure the solution is usable and sustainable. For this purpose, we collaborate closely with experts from the fields of technology acceptance and usability. Thanks to our heterogeneous teams, the interdisciplinary perspective can have a supportive effect in order to strengthen user-friendliness and thereby actual usage of the Service System Platform.

## 5. Conclusions

In conclusion, the developed Service Platform Prototype together with the Pre-built Packages represent an essential step forward in managing and facilitating medical research studies, with an emphasis on data harmonization, resource efficiency, data security, and collaborative effectiveness.

## Supporting information

observation_vital_parameter_loinc_code

Structural Mapping Guideline MII CDS schema to FHIR

## 7. Author Contributions

KH: Conceptualization, Investigation, Methodology, Software, Visualization, Project administration, Writing – original draft, Writing – review & editing; IN: Conceptualization, Methodology, Software, Project administration, Writing – review & editing; YP: Resources, Software, Writing – review & editing; EH: Resources, Software, Writing – review & editing; DB: Resources, Software, Writing – review & editing; CK: Software, Writing – review & editing; MG: Resources, Writing – review & editing; MB: Validation, Writing – review & editing; FN: Writing – review & editing; RG: Writing – review & editing; SG: Resources, Writing – review & editing; AS: Resources, Writing – review & editing; FB: Conceptualization, Funding acquisition, Resources, Writing – review & editing; IR: Resources, Writing – review & editing; MW: Supervision, Writing – review & editing; JW: Writing – review & editing; MS: Funding acquisition, Supervision, Writing – review & editing.

## 9. Acknowledgments

Not applicable.

## 11. Supplementary Material

S1 File: Example table for the semantic mapping of vital parameter: observation_vital_parameter_loinc_code.csv

(CSV)

S2 File: Structural Mapping Guideline MII CDS schema to FHIR (Excel)

## 12. Data Availability Statement

The source code of the pre-built packages “ResearchData-to-FHIR v2.2.0”, “FHIR-to-OMOP v1.1.0”, “Addons v1.0.0” and “MiHUBx Service Platform v1.0.0” can be downloaded from the GitLab repository at https://gitlab.ukdd.de/pub/mihubx. These repositories include the test datasets, developer documentation and the initial installation instructions.

